# Giving birth under hospital visitor restrictions: Heightened acute stress in childbirth in COVID-19 positive women

**DOI:** 10.1101/2020.11.30.20241026

**Authors:** Gus A Mayopoulos, Tsachi Ein-Dor, Kevin G Li, Sabrina J Chan, Sharon Dekel

## Abstract

As the novel coronavirus (COVID-19) has spread globally, a significant portion of women have undergone childbirth while possibly infected with the virus and also under social isolation due to hospital visitor restrictions. Emerging studies examined birth outcomes in COVID-19 positive women, but knowledge of the psychological experience of childbirth remains lacking. This study survey concerning childbirth and mental health launched during the first wave of the pandemic in the US. Women reporting confirmed/suspected COVID-19 during childbirth were matched on various background factors with women reporting COVID-19 negative. We found higher prevalence of clinically significant acute stress in birth in COVID-19 positive women. This group was 11 times as likely to have no visitors than matched controls and reported higher levels of pain in delivery, lower newborn weights, and more infant admission to neonatal intensive care units. Visitor restrictions were associated with these birth outcomes. COVID-19 positive women with no visitors were 6 times as likely to report clinical acute stress in birth than COVID-19 positive women with visitors. The findings underscore increased risk for childbirth-induced psychological morbidity in COVID-19-affected populations. As hospitals continue to revise policies concerning visitor restrictions, attention to the wellbeing of new mothers is warranted.

## Introduction

The coronavirus (COVID-19) pandemic’s immense scope and duration has made clear the urgent need to better understand the virus’ physical and psychological impacts on vulnerable populations. From a generational health perspective, perhaps no population’s experience is more critical to understand and safeguard than that of delivering mothers. In the midst of a global public health crisis characterized by a potentially lethal and highly infectious virus, many non-emergency hospital-based health procedures were postponed. Nevertheless, delivering mothers all over the globe were among the very few populations that continued to be treated in hospital settings.

A significant portion of them underwent childbirth when they were suspected or confirmed of the novel coronavirus disease. Some may have experienced mild but also severe physical symptoms [1] such as fever, lymphocytopenia, and elevated C-reactive protein [2, 3] in accords with reports that pregnancy may result in acute immune changes and cause a viral illness to be more severe [4]. Many mothers, even if asymptomatic, may have experienced heightened emotional distress surrounding the concern that they might be contracting the virus in the hospital or transmitting it to their infant [5]. Labor and delivery present physical and psychological challenges during normalcy, therefore it is critical to understand the effect of COVID-19 on delivering mothers. At present, the impact of being COVID-19 positive, confirmed or suspected, on the childbirth experience and maternal and neonatal outcomes remains not fully clear.

Emerging studies have focused largely on obstetrical and neonatal correlates of COVID-19 infection status. A body of research suggests that there is increased risk for adverse outcomes in pregnant women with COVID-19. In a recent systematic review of nine studies in China, the incidence of preterm births, low birth weight, C-section, and NICU admission were found to be higher in COVID-19 positive cases than in the general population [6]. Likewise, in a second review of 41 cases, higher rates of preterm birth and NICU admission were found in positive pregnancies in comparison with non-positive [7], although other studies did not find differences between affected and non-affected women in maternal and neonatal outcomes [8, 9]. The inclusion of studies with small samples and inappropriate control groups in the reviews above may limit findings and interpretations.

An important issue to consider is the subjective experience of childbirth and potential heightened psychological adversity in birth for COVID-19-positive laboring women. Although childbirth is typically considered a happy event, a significant proportion of new mothers report their delivery as highly stressful in samples assessed before the pandemic [10, 11] and childbirth pain as the most agonizing of painful experiences [12]. To the best of our knowledge, no study has examined how giving birth while potentially being ill with the virus may amplify acute traumatic stress responses to childbirth.

A salient factor that may influence the childbirth of COVID-19-affected mothers concerns the unique stressor of the pandemic: social isolation. With the goal of reducing infectious exposures to visitors, other patients, the community, and healthcare teams, and in the wake of uncertain and rapidly evolving situations, policies restricting visitors have been implemented to lower the number of people in hospitals. Accordingly, mothers who contracted COVID-19 have faced drastic restrictions on maternal visitors and supportive companions in the delivery room and during the postpartum stay. A significant number of COVID-19-affected mothers may have even experienced childbirth without the emotional support provided by having close friends or family in the room with them [13]. They may also have gone without visitors while being separated from their newborn to reduce additional transmission risks [8]. Continuous support in labor and delivery can improve obstetrical and neonatal outcomes and reduce negative birth perceptions, which has been documented before the pandemic [14, 15]. It may further buffer against traumatic stress in response to birth [10].

As the obstetric health risks and benefits in the face of a still poorly understood virus remain unclear [16] and hospitals across the United States continue to evaluate and adjust their visitor policies in light of recommendations from the Centers for Disease Control and Prevention (CDC) and the American College of Obstetricians and Gynecologists (ACOG), a better understanding of the psychological childbirth experiences in COVID-19 vulnerable mothers, such as those being suspected or confirmed of infection, is warranted. In the writing of this work, visitor prohibitions have been largely lifted in maternity wards. However, those delivering who test positive for COVID-19 may still face social isolation and not be allowed any visitors during their entire hospital stay, underlining the importance of relevant research.

To this end, we studied a large sample of women who recently gave birth when COVID-19 was prevalent in the United States, among them 68 women reported suspected or confirmed COVID-19 positive. We matched this group on a wide range of background factors to 68 women who gave birth in the outbreak of the pandemic but were negative for COVID-19. There have been no studies to date that use a comprehensive matched-group analysis that could allow for better understanding of the contribution of COVID-19 positivity to childbirth outcomes while controlling for background factors that increase perinatal adversity. We examined whether being COVID-19 positive is associated with stressful psychological experiences of birth as well as obstetrical and neonatal outcomes and whether having no visitors during delivery hospitalization stay was associated with these outcomes.

## Methods

### Participants

This study is part of a research project that was launched on April 2nd, 2020, in the midst of the COVID-19 pandemic in the United States, with the overarching goal of understanding the impact of COVID-19 on childbirth and maternal mental health. Women who had given birth in the last six months were recruited through announcements on our hospital’s research study platform as well as via social media and postpartum professional communities; they were asked to complete an anonymous survey and were informed that by agreeing to complete the survey they are implying their consent to participate in the study. Therefore, all subjects who took the survey consented to the study. Partners Healthcare (Mass General Brigham) Human Research Committee approved the study measures and procedures and granted exemption for this study and the study was carried out in accordance with the approved protocol. The sample in this study was derived from 2,417 women who gave birth since COVID-19 was prevalent in their communities and provided the childbirth date; they were on average two months postpartum. We identified 68 women who reported being COVID-19 positive, suspected or confirmed, during pregnancy and/or childbirth. We then identified a matched control group of 68 women who reported being COVID-19 negative. The groups were matched on demographic factors, primiparity, prior trauma and childbirth history, and prior mental health.

In this sample of a total of 136 postpartum women, the vast majority delivered a healthy baby at term (86.8%), had a vaginal delivery (71.3%), and around half (50.7%) were primiparas. The average age of participants was 32 years old. The majority were married (89%), had at least middle-class income (i.e., $100,000 per year, 66.2%), were employed (72.1%), and had at least a college degree (83.1%). Participants resided in the United States (80.0%), in Canada (4.4%), Europe (2.9%), Central/South America (2.9%), Asia (2.9%), and 2.2% in the Caribbean and Middle East. Four participants (2.9%) did not report their geographic location.

### Measures

Acute stress responses to childbirth were assessed with the commonly used Peritraumatic Distress Inventory (PDI) [17]. The PDI is a 13-item self-report with good psychometric properties. It assesses negative emotional responses (e.g., “I felt helpless”; “I thought I might die”) experienced during and/or immediately after a specified traumatic event on a 0 (not at all) to 4 (extremely true) scale. In this study, participants rated their responses in regard to their recent childbirth experience. The PDI has been used to assess acute childbirth-related stress in postpartum samples [18]. To define clinically significant acute stress response symptoms, we used the suggested cutoff of 17 [19]. Reliability in the current study was high (α = 0.91).

Obstetrical and infant factors concerning recent childbirth were measured with respect to gestational age, medical complications in labor and delivery (yes vs. no), degree of pain in labor and delivery (assessed on a 5-point Likert Scale), sleep deprivation (defined as less than six hours of sleep on the night before childbirth), use of pain medication (yes vs. no), use of induction medication (yes vs. no), and mode of delivery. Additionally, we measured newborn weight (lbs.), newborn biological sex, neonatal intensive care unit (NICU) admission of newborn, skin-to-skin contact after delivery (yes vs. no), rooming-in (yes vs. no) and breastfeeding habits (exclusive, mixed, stopped, or never breastfeeding offered).

COVID-19-related restrictions were assessed in regard to visitor policy. Participants were asked whether there were “any visitor restrictions during your hospital stay?”. Response options (no visitors, one visitor, no restrictions) were classified as “No visitors” versus “Other”. We also asked participants whether they were separated from their infant.

Background factors pertaining to demographics, prior mental health, and traumatic exposure were used for creating matching study groups. Theses variables included maternal age, education level, marital and employment status, income, race/ethnicity, and primiparity. We also assessed history of mental health problems (i.e., depression, postpartum depression, anxiety, posttraumatic stress disorder), and the number of prior traumatic events (happened or witnessed) with the commonly used Life Events Checklist for DSM-5 (LEC-5) [20] (α = 0.91); as well as stressors in previous pregnancies (defined as miscarriage, stillbirth, or premature delivery).

### Data Analysis

To create matched groups who share similar background characteristics between COVID-19 positive and negative women, we conducted a propensity score matching procedure using SPSS PS module [21] (see detailed variables listed in the Methods). The estimation algorithm was logistic regression, the matching algorithm was nearest neighbor matching with caliper of 0.2 as recommended by Austin [22]. An overall balance test [23] was performed to estimate the balance in the matching process.

Following the matching procedure, we compared groups in obstetrical factors (sleep deprivation, pain in labor and birth, birth complications, medication for induction and pain, and mode of delivery), infant-related factors (gestational age, NICU admission, weight and sex, breastfeeding, rooming in, and skin-to-skin), COVID-19-related restrictions (separation from newborn and lack of visitors during delivery hospitalization), and psychological experience of birth, namely, acute stress responses in birth. Kolmogorov-Smirnov and Shapiro-Wilk tests were conducted to assess normality in quantitative outcome scores. Based on these tests, Mann-Whitney U tests were performed to estimate differences between study groups in quantitative measures, and chi-square test for independence of measures (with Fisher’s exact estimation of significance) for estimating differences in categorical measures. Finally, we conducted Kruskal-Wallis tests and chi-square test for independence of measures (with Fisher’s exact estimation of significance) to examine whether no visitors may account for the differences between COVID-19 positive and negative women.

## Results

### Group matching

An overall balance test [23] indicated that the balance of the matching was high, *χ*^*2*^_(27)_ = 18.21, *p* = .90, such that each group comprised 68 women.

### COVID-19 positive birth-related outcomes

Percentages of birth-related outcomes are presented in Table 1; mean differences are presented in Table 2.

**Table 1.** Differences in the percentage of birth-related outcomes

| | COVID-19 | | | | $\chi^2$ | OR (95% CI) |
| --- | --- | --- | --- | --- | --- | --- |
|  | Positive |  | Negative |  |  |  |
|  | % | n | % | n |  |  |
| Sleep deprivation | 70.6 | 48 | 64.7 | 44 | 0.54 | 1.31 (0.64, 2.69) |
| Complications | 27.9 | 19 | 19.1 | 13 | 1.47 | 1.64 (0.73, 3.66) |
| Medication for induction | 47.1 | 32 | 47.1 | 32 | 0.00 | 1.00 (0.51, 1.96) |
| Medication for pain | 73.5 | 50 | 80.9 | 55 | 1.05 | 0.66 (0.29, 1.48) |
| Mode of delivery |  |  |  |  | 3.74 |  |
| Natural | 22.1 | 15 | 17.6 | 12 |  | 1.32 (0.57, 3.08) |
| Vaginal | 52.9 | 36 | 50.0 | 34 |  | 1.13 (0.57, 2.20) |
| Assisted | 7.4 | 5 | 2.9 | 2 |  | 2.61 (0.49, 13.99) |
| Planned Cesarean | 8.8 | 6 | 16.2 | 11 |  | 0.50 (0.17, 1.44) |
| Unplanned Cesarean | 8.8 | 6 | 13.2 | 9 |  | 0.63 (0.21, 1.89) |
| NICU admission | 17.6 | 18 | 8.8 | 6 | 2.31 | 3.72** (1.37, 10.07) |
| Infant's sex (boys) | 46.3 | 31 | 56.3 | 36 | 1.31 | 0.74 (0.38, 1.46) |
| Breastfeeding |  |  |  |  | 1.23 |  |
| Exclusive | 67.6 | 46 | 66.2 | 45 |  | 1.06 (0.52, 2.18) |
| Breastfeeding + supplement | 19.1 | 13 | 22.1 | 15 |  | 0.84 (0.36, 1.92) |
| Stopped | 8.8 | 6 | 10.3 | 7 |  | 0.84 (0.27, 2.65) |
| No breastfeeding | 4.4 | 3 | 1.5 | 1 |  | 3.09 (0.31, 30.50) |
| Rooming in | 85.3 | 58 | 97.1 | 66 | 5.85* | 0.18* (0.04, 0.84) |
| Skin-to-skin contact | 79.4 | 54 | 88.2 | 60 | 1.95 | 0.51 (0.20, 1.32) |
| Separation from infant | 17.6 | 12 | 0.0 | 0 | 13.16*** | 30.31*** (1.76, 523.26) |
| No visitors in hospital stay | 25 | 17 | 2.9 | 2 | 13.77*** | 11.0*** (2.42, 49.80) |
| Acute stress response in childbirth | 47.1 | 32 | 29.4 | 20 | 4.48* | 2.13* (1.05, 4.32) |

Note. \* $p < .05$ , \*\* $p < .01$ , \*\*\* $p < .001$ ; OR = Odd ratios, 95% CI = 95% confidence interval.

**Table 2.**
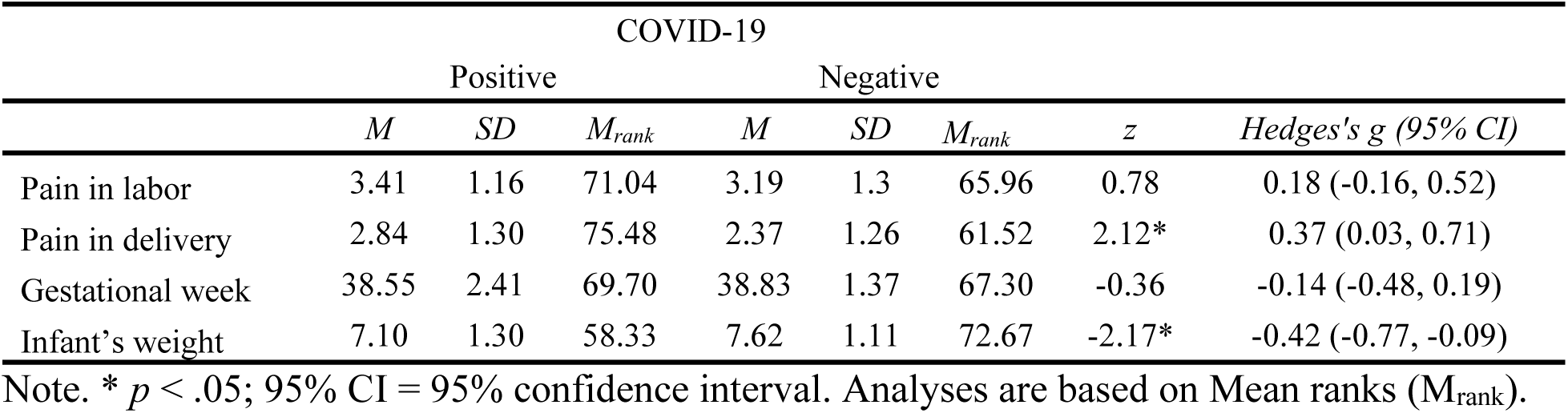
Differences in the mean level of birth-related outcomes

#### Obstetrical-related factors

COVID-19 positive women reported significantly greater pain in delivery (weak-to-moderate in effect size) than COVID-19 negative women. No group differences were found in sleep deprivation, pain in labor, medical complications, medication for induction and/or pain, or mode of delivery.

#### Infant-related factors

A higher percentage of babies of COVID-19 positive women were admitted to the newborn intensive unit of care (*OR* = 3.72). In addition, COVID-19 positive women gave birth to infants with lower (yet normal) weights than COVID-19 negative women. Fewer COVID-19 positive women were with their newborn in the room during their stay at the hospital (*OR* = 0.18) than COVID-19 negative women. No group differences were found in gestational age, infant sex, breastfeeding, and skin-to-skin contact.

#### COVID-19 restrictions

More COVID-19 positive women were separated from their newborns and had no visitors during hospitalization stay than COVID-19 negative women.

#### Psychological experience of birth

More COVID-19 positive women had clinical levels of acute stress response to birth than COVID-19 negative women.

### Does lack of visitors account for the differences between the COVID-19 positive and negative groups?

To examine whether no visitors during delivery hospitalization account for the differences between the COVID-19 positive and negative groups (i.e. in pain in delivery, NICU admission, infant weight, rooming in, and acute stress response to birth), we compared COVID-19 negative women who had visitors (*n* = 66), with COVID-19 positive women who had visitors (*n* = 51) and those with no visitors (*n* = 17). Percentages are presented in Table 3; mean differences are presented in Table 4.

**Table 3.** Differences in the percentage of birth-related outcomes as a function of COVID and no visitors during delivery hospitalization

| | COVID-19 negative | | Study group<br>COVID-19 positive/ visitors | | COVID-19 positive/ no visitors | | $\chi^2$ | OR (95% CI) |
| --- | --- | --- | --- | --- | --- | --- | --- | --- |
|  | % | *n* | % | *n* | % | *n* |  |  |
| NICU admission | 9.1 | 6 | 9.8 | 5 | 41.2 | 7 | 12.19** | 6.16 (1.61, 25.67)<sup>1</sup><br>6.72(1.84, 25.91)<sup>2</sup> |
| Rooming in | 97.0 | 64 | 94.1 | 48 | 58.8 | 10 | 25/08*** | 0.10 (0.02, 0.42)<sup>1</sup><br>0.05(0.01, 0.25)<sup>2</sup> |
| Acute stress response | 22.7 | 15 | 54.9 | 28 | 88.2 | 15 | 28.98*** | 5.70 (1.39, 42.44)<sup>1</sup><br>22.98 (5.59, 172.33)<sup>2</sup> |

Note. \* $p < .05$ , \*\* $p < .01$ , \*\*\* $p < .001$ ; OR = Odd ratios, 95% CI = 95% confidence interval. <sup>1</sup> = COVID-19 positive / visitors vs. COVID-19 positive / no visitors; <sup>2</sup> = COVID-19 negative vs. COVID-19 positive / no visitors

**Table 4.** Differences in the mean level of birth-related outcomes as a function of COVID and lack of visitors

| | COVID-19 negative | | | Study group<br>COVID-19 positive / visitors | | | COVID-19 positive / no visitors | | | Kruskal–Wallis *H* | $\eta_p^2$<br>(95% CI) |
| --- | --- | --- | --- | --- | --- | --- | --- | --- | --- | --- | --- |
|  | *M* | *SD* | *M<sub>rank</sub>* | *M* | *SD* | *M<sub>rank</sub>* | *M* | *SD* | *M<sub>rank</sub>* |  |  |
| Pain in delivery | 2.38 | 1.26 | 60.68 | 2.65 | 1.35 | 68.20 | 3.41 | 0.94 | 91.88 | 9.21** | 0.06 (0.01, 0.13) |
| Infant's weight | 7.60 | 1.12 | 71.20 | 7.37 | 0.91 | 62.99 | 6.20 | 1.92 | 41.40 | 7.96* | 0.12 (0.04, 0.20) |

Note. \* $p < .05$ ; 95% CI = 95% confidence interval. Analyses are based on Mean ranks ( $M_{rank}$ ).

The analyses indicated that COVID-19 positive women who had no visitors reported significantly greater pain in delivery (see Figure 1) and delivered infants with lower weights (see Figure 2). In addition, their infants were more likely to get admitted to the NICU and less likely to be in the same room with their mothers during the hospital stay. Finally, COVID-19 positive women with no visitors had much higher prevalence of acute stress responses at a clinical level (see Figure 3).

**Figure 1.**
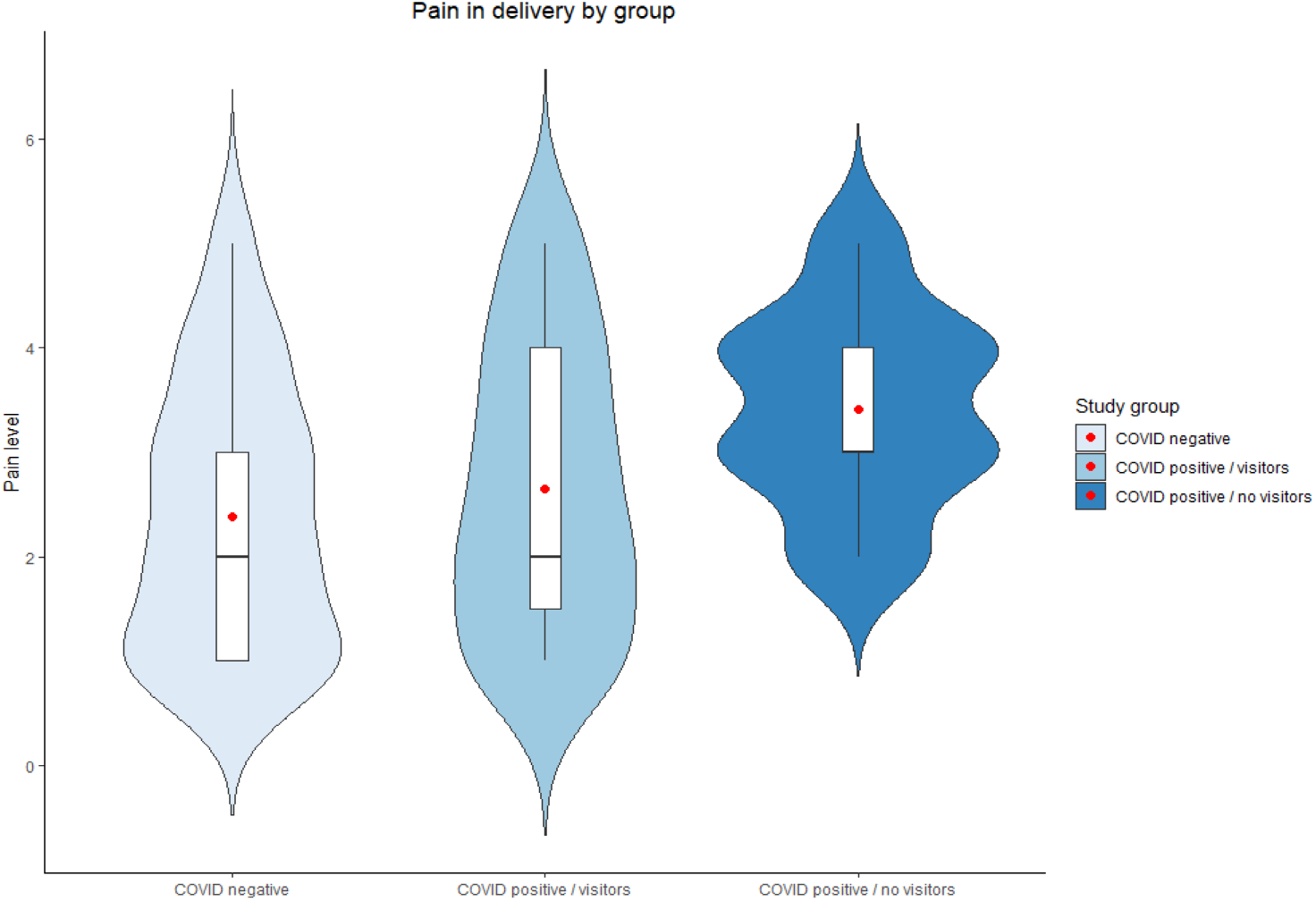
Pain in delivery by study group. Red dots represent the mean.

**Figure 2.**
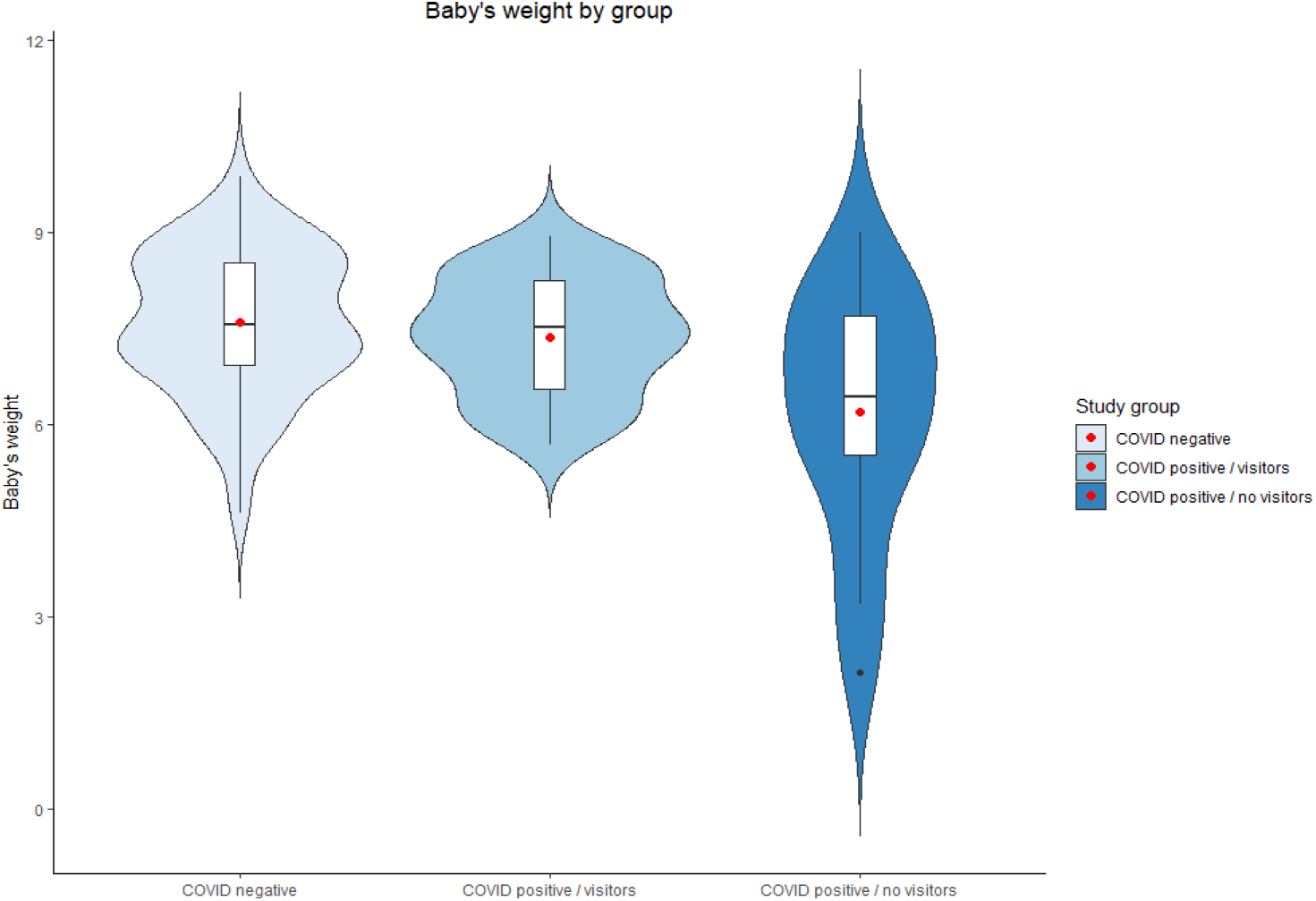
Infant’s weight by study group. Red dots represent the mean.

**Figure 3.**
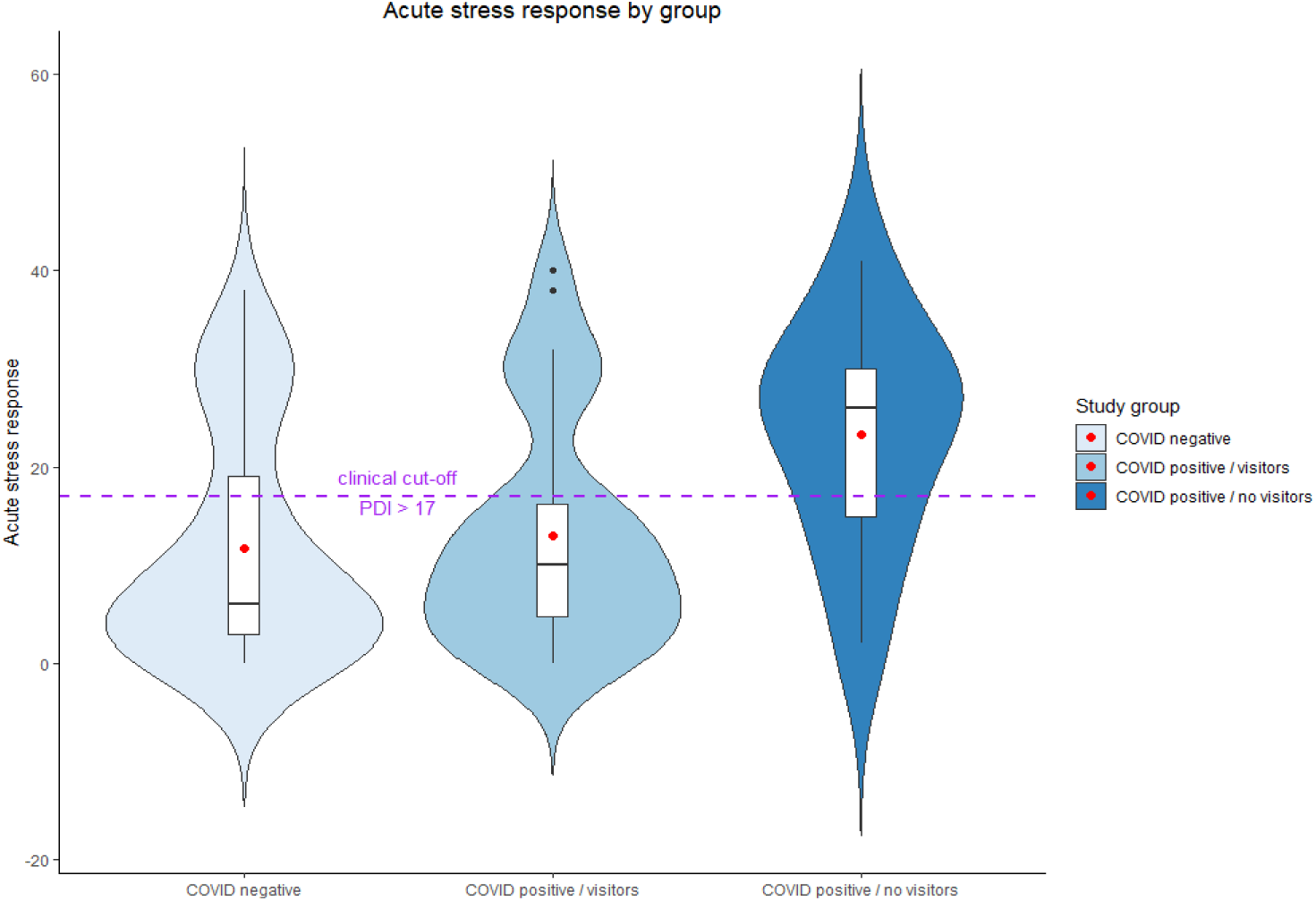
Acute stress response to childbirth by study group. Red dots represent the mean; dashed purple line represent the clinical cutoff.

## Discussion

The COVID-19 pandemic offers a rare opportunity to examine the experience of childbirth under stressful conditions such as social isolation. As infectious disease outbreaks continue, it is critical that we generate new knowledge to inform preparations and guidelines of perinatal care during these outbreaks.

Our study sought to examine the childbirth experiences of women who had delivered during peak infection rates of the pandemic and were suspected or confirmed to have contracted COVID-19, a population likely to have undergo labor and delivery while potentially being acutely ill and at the same time subject to drastic hospital restrictions concerning social isolation. We compared women who reported being COVID-19 positive, suspected or confirmed, to women who reported not having contracted the virus but who were similar on a range of background factors such as demographics, prior mental health, and even trauma history. This rigorous matched-control group approach has not been implemented in previous studies and allows for the generation of knowledge on the potential adverse influence of COVID*-*19 infection status of birth outcomes while controlling for background factors that are associated with COVID-19 infection and negative maternal outcomes.

The main study findings show that nearly 50% of suspected or confirmed COVID-19 positive women reported clinically significant acute stress symptoms in birth. They were as much as two times more likely to experience acute stress than non-affected women and to perceive higher degrees of pain in childbirth even though no differences were found in factors such as obstetrical complications, medication for pain, or delivery mode between COVID-19 positive and negative cases. These findings underscore how childbirth can become a traumatic experience and evoke an acute stress response for women with the novel coronavirus.

We further document increased exposure to salient social stressors surrounding childbirth in affected women. As might be expected, the results reveal that hospital policies enforcing visitor restrictions were frequently implemented with delivering women suspected or confirmed of COVID-19 infection. As much as 25% of COVID-19 positive women had no visitors during their delivery hospitalization stay. This group was 11 times as likely not to be permitted a support person to accompany them than women negative for COVID-19. COVID-19 positive women were also much more likely to experience physical separation from their newborn. In accord with previous studies [7], the newborns were nearly four times as likely to be admitted to the NICU. In the writing of this manuscript, the CDC has updated its guidance and currently recommends rooming-in for a COVID-positive mother and her newborn and acknowledges that the decision should be determined by the family.

Our findings reveal that social isolation surrounding childbirth may increase risk for maternal morbidity. We found heightened clinically significant acute stress in COVID-19 positive women who had no visitors. They were 6 times as likely to report acute stress symptoms than COVID-19 affected women who were permitted visitors during their delivery hospitalization. We also found that COVID-19 positive women who did not have a support person experienced greater pain in delivery, delivered newborns with lower weight, and had elevated NICU admission rates. These findings accord with the evidence of emotional comfort and support in birth being associated with improved birth outcomes [14], and suggest how a diminished sense of support may increase maternal stress and subsequent adversity. Psychological traumatic morbidity in birth has been shown to result in maternal mental illness during the postpartum period based on pre-COVID samples [24, 25] and has also been documented in women who gave birth since the pandemic [26].

This study’s findings may be useful in informing clinical policies during the COVID-19 pandemic. While much attention has been paid to the physical symptoms in mothers with COVID-19, our study emphasizes the importance of considering mothers’ psychological wellness. The findings suggest that increased awareness should be given in labor and delivery and postpartum units to the psychological symptoms surrounding childbirth that may arise in women who are sick with or suspected of having the virus; additionally, the potential emotional liability of not permitting a support person during hospitalization should be noted. While routine screening for traumatic childbirth does not exist in postpartum hospital units, our study suggests that assessment of acute stress responses in delivering mothers who are COVID-19 positive is warranted. Ongoing monitoring of mental health symptoms in this high risk group after hospital discharge is important as those with stable symptoms in accord with routine care are likely to be quickly discharged and face social isolation during the postpartum period, which is considered a time of heightened psychological vulnerability [27, 28].

Shortcomings of this study include reliance on anonymous self-report measures that allowed for conducting a study swiftly during the initial heights of the pandemic but not for inclusion of patients’ medical records. We rely on respondents accurately reporting their COVID-19 infection status, and their receiving accurate information from COVID-19 testing protocols at the hospitals where they delivered. Additionally, we do not have information on the severity of respondents’ COVID-19 symptoms, only their infection status. We cannot rule out that acutely ill women were those not permitted visitors. Also, while we used a well-validated measure to assess acute stress which has shown good correspondence with clinician assessments, we did not include diagnostic measures. Retrospective assessments could be prone to recall bias and hence the importance of the use of matched controls. This convenient internet sample introduces a bias towards women from a certain socioeconomic class.

## Conclusion

In conclusion, we find that confirmed or suspected COVID-19 positive women experience increased psychological morbidity surrounding childbirth compared to delivering women without COVID-19. We find that COVID-19 positive women experience increased levels of pain during delivery and give birth to newborns of lower weight which are more likely to be separated from their mothers and sent to the NICU. This increased adversity appears especially heightened in cases where a support person is not allowed in the maternity unit. As hospitals around the world continue to update their delivery protocols for COVID-19 positive women and determine risk and benefits of visitor restriction policies, more research is needed to optimize maternal care during these unprecedented times.

## Data Availability

The dataset is available on Dryad. It is currently set to private for peer review but can be viewed at the link below.

https://datadryad.org/stash/share/zWK2HL8u-0wzNj1ilX6jfqNLYL-q4kkSnr6ketOpBnk

## Acknowledgements

We would like to thank Ms. Gabriella Dishy for her assistance in building the web survey and Ms. Rasvitha Nandru and Ms. Aruni Ahilan for their assistance in the recruitment of study participants.

## Author Contributions

GM wrote the manuscript. TE conducted the statistical analysis and wrote the Methods. KL contributed to the writing of the manuscript. SC contributed to the data collection and the editing of the manuscript. SD is the principal investigator of the larger project. She designed the current study, collected the data, and supervised the manuscript preparation. All authors read and approved the final manuscript.

## Additional Information

The authors declare no competing interests.

## References

1. Andrikopoulou, M., et al. Symptoms and critical illness among obstetric patients with coronavirus disease 2019 (COVID-19) infection. Obstet. Gynecol. 136, 291–299 (2020).

2. Liu, D., et al. Pregnancy and perinatal outcomes of women with coronavirus disease (COVID-19) pneumonia: a preliminary analysis. AJR Am. J. Roentgenol. 215, 127–132 (2020).

3. Zaigham, M. & Andersson, O. Maternal and perinatal outcomes with COVID-19: a systematic review of 108 pregnancies. Acta Obstet. Gynecol. Scand. 99, 823–829 (2020).

4. Silasi, M., et al. Viral infections during pregnancy. Am. J. Reprod. Immunol. 73, 199–213 (2015).

5. Panahi, L., Amiri, M., & Pouy, S. Risks of novel coronavirus disease (COVID-19) in pregnancy; a narrative review. Arch. Acad. Emerg. Med. 8, 34 (2020).

6. Smith, V., et al. Maternal and neonatal outcomes associated with COVID-19 infection: a systematic review. PLoS One. 10.1371/journal.pone.0234187 (2020).

7. Di Mascio, D., et al. Outcome of coronavirus spectrum infections (SARS, MERS, COVID 1-19) during pregnancy: a systematic review and meta-analysis. Am. J. Obstet. Gynecol. MFM. 2, 100107; 10.1016/j.ajogmf.2020.100107 (2020).

8. Griffin, I., et al. The impact of COVID-19 infection on labor and delivery, newborn nursery, and neonatal intensive care unit: prospective observational data from a single hospital system. Am. J. Perinatol. 37, 1022–1030 (2020).

9. Nayak, A. H., et al. Impact of the coronavirus infection in pregnancy: a preliminary study of 141 patients. J. Obstet. Gynaecol. India. 2020;70(4):256–61.

10. Dekel, S., Stuebe, C., & Dishy, G. A. Childbirth induced posttraumatic stress syndrome: a systematic review of prevalence and risk factors. Front. Psychol. 8, 560 (2017).

11. Yildiz, P. D., Ayers, S., & Phillips, L. The prevalence of posttraumatic stress disorder in pregnancy and after birth: a systematic review and meta-analysis. J. Affect Disord. 208, 634–645 (2017).

12. Pirdel, M. & Pirdel, L. Perceived environmental stressors and pain perception during labor among primiparous and multiparous women. J. Reprod. Infertil. 10, 217–223 (2009).

13. Ecker, J. L. & Minkoff, H. L. Laboring alone? Brief thoughts on ethics and practical answers during the coronavirus disease 2019 pandemic. Am. J. Obstet. Gynecol. MFM. 2, 100141; 10.1016/j.ajogmf.2020.100141 (2020).

14. Tani, F. & Castagna, V. Maternal social support, quality of birth experience, and post-partum depression in primiparous women. J. Matern. Fetal Neonatal Med. 30, 689–692 (2017).

15. WHO: Why having a companion during labor and childbirth may be better for you. 2019.

16. Arora, K. S., Mauch, J. T., & Gibson, K.S. Labor and delivery visitor policies during the COVID-19 pandemic: balancing risks and benefits. JAMA. 323, 2468–2469 (2020).

17. Brunet, A., et al. The Peritraumatic Distress Inventory: a proposed measure of PTSD criterion A2. Am. J. Psychiatry. 158, 1480–1485 (2001).

18. Dekel, S., Ein-Dor T., Dishy, G. A., & Mayopoulos, P. A. Beyond postpartum depression: posttraumatic stress-depressive response following childbirth. Arch. Womens Ment. Health. 23, 557–564 (2020).

19. Nishi, D., et al. Peritraumatic distress inventory as a predictor of post-traumatic stress disorder after a severe motor vehicle accident. Psychiatry Clin. Neurosci. 64, 149–156 (2010).

20. Weathers, F., et al. The life events checklist for DSM-5 (LEC-5). National Center for PTSD www.ptsd.va.gov (2013).

21. Thoemmes, F. Propensity score matching in SPSS. Preprint at https://arxiv.org/abs/1201.6385 (2012).

22. Austin, P. C. Optimal caliper widths for propensity-score matching when estimating differences in means and differences in proportions in observational studies. Pharm. Stat. 10, 150–161 (2011).

23. Hansen, B. B. & Bowers, J. Covariate balance in simple, stratified and clustered comparative studies. Statist. Sci. 23, 219–236 (2008).

24. Grekin, R., O’Hara, M. W., & Brock, R. L. A model of risk for perinatal posttraumatic stress symptoms. Arch. Womens Ment. Health. 10.1007/s00737-020-01068-2 (2020).

25. Chan, S. J., et al. Risk factors for developing posttraumatic stress disorder following childbirth. Psychiatry Res. 290, 113090; 10.1016/j.psychres.2020.113090 (2020).

26. Mayopoulos, G. A., et al. COVID-19 is associated with traumatic childbirth and subsequent mother-infant bonding problems. Manuscript submitted for publication (2020).

27. Putnick, D. L., et al. Trajectories of Maternal Postpartum Depressive Symptoms. Pediatrics. 146, 20200857; 10.1542/peds.2020-0857 (2020).

28. Dekel, S., et al. Delivery mode is associated with maternal mental health following childbirth. Arch. Womens Ment. Health. 22, 817–824 (2019).

